# Prone Positioning in a North American Cohort of Hypoxemic Patients on Mechanical Ventilation

**DOI:** 10.1101/2025.09.03.25334942

**Authors:** Anna K Barker, Akihiko Nishimura, Mark Nuppnau, Kevin G. Buell, Patrick G Lyons, Wan-Ting Liao, Brenna Park-Egan, Benjamin E Schmid, Nicholas E Ingraham, Vaishvik Chaudhari, Catherine A Gao, Alexander C Ortiz, Gary E Weissman, Kaveri Chhikara, Juan C Rojas, Andre CKB Amaral, William F Parker, Theodore J Iwashyna, David N Hager, Michael W Sjoding, Chad H Hochberg, CLIF Consortium

## Abstract

**Objective:** Use of prone positioning increased among mechanically ventilated patients during the COVID-19 pandemic, but it is unknown whether implementation of this life-saving intervention was sustained. Thus, we aimed to evaluate peri-pandemic trends in proning use.

**Design:** We conducted a retrospective cohort study of proning use among mechanically ventilated adults, with proning rates compared across pre-pandemic (1/2018-2/2020), pandemic (3/2020-2/2022), and post-pandemic (3/2022-12/2024) periods.

**Setting:** 37 North American hospitals

**Patients:** Mechanically ventilated patients with persistent moderate-to-severe hypoxemia (PaO_2_/F_i_O_2_ ≤150 mmHg, F_i_O_2_ ≥0.6, and positive end-expiratory pressure ≥5 cmH_2_O).

**Intervention:** Proning within 12 hours of meeting study hypoxemia criteria.

**Measurements and Main Results:** Among 5,760 proning-eligible patients, 1,737 (30.2%) received proning: 8.0% pre-pandemic, 44.6% pandemic, and 19.9% post-pandemic. The adjusted odds ratio (OR) for proning during pandemic versus pre-pandemic periods was 8.25 (95% Confidence Interval (CI): 6.35-10.70) and pandemic versus post-pandemic, 2.76 (95% CI: 1.83-4.17). Proning varied widely by hospital and was quantified with the median odds ratio (median change in odds of proning an identical patient admitted at a lower versus higher proning hospital) of 2.54 (95% Credible Interval (CrI): 1.75-4.58) pre-pandemic, 2.33 (95% CrI: 1.92-3.04) pandemic, and 2.58 (95% CrI: 1.99-3.73) post-pandemic. Pandemic-period patients with SARS-CoV2 were proned more than those without (OR: 4.55, [95% CI: 3.85-5.56]), but pandemic-period patients without SARS-CoV2 were still proned more than pre-pandemic (OR: 3.87, [95% CI: 2.92-5.13]) or post-pandemic patients (OR: 1.37, [95% CI: 1.03-1.83]).

**Conclusions:** In a North American cohort of proning-eligible patients, proning increased during the pandemic and then declined. Interventions that improve implementation of this life-saving treatment are urgently needed.

## Introduction

Acute respiratory distress syndrome (ARDS) is common in intensive care units (ICUs), and yields high morbidity and mortality rates approaching 50% for patients with moderate-to-severe ARDS (1). Prone positioning is one of the few life-saving guideline-recommended interventions for ARDS, but has historically been underused. Prior to the COVID-19 pandemic less than one-third of eligible patients received proning (1–4). While proning rates increased during the pandemic, it is unknown whether this improved implementation has been sustained (5–9).

Given its historical underuse, understanding temporal trends and sources of variation in proning rates is critical for determining current implementation needs. While small single-health system studies suggest that proning use may have declined back towards pre-pandemic levels (5, 7), practices vary considerably across institutions and conclusions drawn from single centers may not be generalizable (10). If the decline in proning use is indeed widespread, this will signal a need for new implementation efforts to increase and sustain use of this life saving intervention.

To understand the use of prone positioning over time, we performed a retrospective cohort study of proning practices across 37 North American hospitals within 9 health systems in the recently developed Common Longitudinal ICU data Format (CLIF) research consortium (11). The study sought to describe proning use before, during, and after the pandemic and to quantify hospital-level variation in proning practices. We hypothesized that there would be consortium-wide increases in proning use during the pandemic, followed by a subsequent decline.

## Materials and Methods

### Data Source

We leveraged electronic health record (EHR) databases at 9 health systems in the CLIF consortium to generate a retrospective cohort of mechanically ventilated adults admitted 1/1/2018-12/31/2024 at 1 Canadian and 36 United States hospitals. CLIF consortium sites maintain granular acute care databases locally in an open-source common data format optimized for federated critical care research (11). Sites can use a custom GPT AI tool to facilitate structuring their local CLIF databases (https://clif-consortium.github.io/website/tools.html). In a privacy-preserving approach, common cohort identification and analytic scripts were developed and run within each local CLIF database. Aggregate data were shared and pooled as described below.

Each health system consulted with their local institutional review board who either provided ethical approved for the study or confirmed that no ethical approval was required (Supplement Table-S1). Given that this study was conducted retrospectively from data obtained for clinical purposes, signed written consent was not obtained. Study reporting conforms to the STROBE guidelines for observational studies (Supplement Table-S2; 15).

### Study Population

The cohort was designed to mirror inclusion criteria from PROSEVA, the landmark randomized controlled trial on proning (13). Because the Berlin ARDS definition can be difficult to identify in clinical practice and research (1, 14), the study pragmatically evaluated patients with acute, persistent moderate-to-severe hypoxemic respiratory failure as a proxy for proning-eligible ARDS. We included adults (>18 years) receiving mechanical ventilation who met the following criteria: 1) PaO_2_/F_i_O_2_≤150 mmHg on F_i_O_2_≥0.6 and positive end-expiratory pressure (PEEP) ≥5 cmH_2_O within 36 hours of intubation and 2) a second qualifying PaO_2_/F_i_O_2_≤150 mmHg, on F_i_O_2_≥0.6 and PEEP≥5 cmH_2_O, 12-24 hours after the first qualifying blood gas. To account for improvement in hypoxemia after proning, patients proned within 24 hours of the initial qualifying blood gas were also included, even without a second qualifying blood gas. We excluded those with: 1) mechanical ventilation initiated outside the local health system, 2) tracheostomy present on the day of intubation, and 3) procedural location 48-hours prior to meeting study criteria, to exclude recent surgical patients with potentially high rates of relative contraindications to proning (13). For patients with multiple hospitalizations meeting criteria, one encounter was randomly selected for inclusion.

### Exposures and Outcomes

Proning use was evaluated during three pre-specified periods based on predominant trends in COVID-19 activity [18]: 1) pre-pandemic (1/1/2018-2/29/2020), 2) pandemic (3/1/2020-2/28/2022), and 3) post-pandemic (3/1/2022-12/31/2024). We selected the pandemic as the reference period for the primary analysis, to enable comparison of practice patterns across consecutive periods (pre-pandemic:pandemic, pandemic:post-pandemic). Six health systems contributed data across all periods, one during pre-pandemic and pandemic periods, and two during pandemic and post-pandemic periods. As a secondary exposure we evaluated the association of hospital type (community versus academic) and hospital ICU capacity (<20, 20-49, and ≥50 total ICU beds), with proning.

The primary outcome was early prone positioning within 12 hours of meeting eligibility criteria (13). Prone positioning was defined similarly across CLIF health systems using position documentation entered in EHR clinical flowsheets (Supplement Table-S3). This method was highly accurate for proning identification and timing compared to manual chart adjudication at two CLIF health systems (5, 8).

### Patient and hospital-level covariates

We extracted patient-level covariates including: age, sex, race, ethnicity, body mass index (BMI), vasopressor use, arterial blood gases, PEEP, F_I_O_2_, and SARS-CoV2 test results. A modified non-respiratory sequential organ failure assessment (SOFA) score was calculated using data within 24 hours prior to meeting eligibility criteria (16) and included elements for: Glasgow Coma and Richmond Agitation-Sedation Scales (17), mean arterial blood pressure, vasopressor use, and bilirubin, platelet, and creatinine levels. If laboratory data were missing, it was imputed as normal (18). The renal component of the modified SOFA score was based on creatinine and did not incorporate urine output. SARS-CoV2 positivity was defined by a positive antigen or polymerase chain reaction test within 4-weeks prior to meeting study criteria. Vasopressor use was categorized into none, norepinephrine equivalent ≤0.1, or >0.1 μg/kg/min (19).

### Statistical Analysis

To describe the cohort, we pooled individual health system data using weighted means and standard deviations (SD) for continuous variables and total count and percentages for categorical variables. Within each site, we constructed multivariable logistic regression models to evaluate the association of early proning with period, adjusting for pre-specified covariates age, sex, BMI, non-respiratory SOFA score, PaO_2_/F_i_O_2_ ratio, and vasopressor use (5, 8, 9). For systems with multiple hospitals, hospital site was included as a fixed effect. In a federated approach, results were pooled using random effects meta-analysis, a strategy that produces results concordant with patient-level analysis (20).

Less than 5% of data elements were missing across the consortium. We performed imputation in one health system where weight was missing for >15% of patients using multiple imputation by chained equations (Supplement Table S4). Weight was not imputed at sites where missingness was 0-15%. Per policy, one health system did not collect any race or ethnicity data.

We performed secondary analyses including: 1) evaluating period trends stratified by SARS-CoV2 positivity and 2) analyses that quantified variability in proning by hospital, hospital type, and hospital ICU capacity. No pre-pandemic patients and few post-pandemic patients had positive SARS-CoV2 tests. Thus, in the SARS-CoV2 secondary analysis, we evaluated early initiation of proning in four pre-specified groups: pre-pandemic, pandemic patients with SARS-CoV2, pandemic patients without SARS-CoV2, and all post-pandemic patients, using the same analytic approaches as the primary analysis.

To quantify variability in proning use across hospitals and evaluate the association between hospital type, ICU capacity, and proning use, we conducted risk-adjusted, across-hospital, and across-period comparisons of predicted proning rates. This was achieved by fitting a Bayesian hierarchical mixed effect model, with effects of hospitals and periods modelled as random (21). For Bayesian priors, see Supplement Table-S5. To fit a federated model without sharing individual-level data, we first fit local logistic regression models with early proning as the outcome and including predictors: age, gender, BMI, vasopressor use, SOFA score and PaO_2_/F_i_O_2._ The fixed effects coefficients were then aggregated with appropriate weights to obtain global coefficient estimates. We used these global estimates to generate the predicted proning rate for each hospital and period while accounting for the fixed effects (e.g., risk-adjusted proning rate). We then constructed final Bayesian hierarchical models using aggregate logistic regression fit to summary-level data by hospital and period and additionally included an offset term for the risk-adjusted log odds of proning at the hospital and period level (generated from the global coefficient estimates), hospital and period random effects, and an additional fixed effect for either hospital type or ICU capacity (22). Given collinearity between hospital type and ICU capacity, we did not fit a model that included both covariates. We excluded hospital and period data in these analyses if fewer than ten patients were available for a hospital during a given study period.

We visualized hospital variation by generating hospital-level predictions from the Bayesian hierarchical models and creating a caterpillar plot. We quantified this variation using median odds ratios stratified by study period (21). The median odds ratio quantifies the median change in odds of proning for a hypothetical identical patient moved from a randomly selected lower to higher proning hospital. Model estimates are reported as median posterior odds ratios with 95% credible intervals generated from posterior draws of 4000 samples.

Finally, we conducted several prespecified sensitivity analyses to evaluate the robustness of our primary results to cohort and outcome definitions and analytic choices for pooling methods. These included: (1) evaluating only patients admitted to the six health systems that contributed data to all three periods, (2) evaluating patients with severe respiratory failure (PaO_2_/F_I_O_2_ <100) upon meeting study eligibility, (3) extending the period for early initiation of proning from 12 to 72 hours, and (4) generating adjusted odds ratios for the primary period estimates using the Bayesian hierarchical approach instead of meta-analysis. All analyses were done with R version 4.3. Statistical code is available on github (https://github.com/Common-Longitudinal-ICU-data-Format/CLIF_Proning_Incidence_Severe_ARF.git). Some results were previously reported at a conference (23).

## Results

Across 9 health systems and 37 hospitals, 5,760 proning-eligible patients on mechanical ventilation were included: 1,195 (20.7%) pre-pandemic, 2,969 (51.5%) pandemic, and 1,596 (27.7%) post-pandemic (Supplement Figure-S1). In the overall cohort, mean age was 59 (SD, 15) years, 2,429 (42.2%) patients were female and 1,716 (29.8%) were positive for SARS-CoV2 (Table 1). The sample included patients at 25 community (1,796 [31.2%] patients) and 12 academic hospitals (3,964 [68.8%] patients).

**Table 1:** Characteristics of Patients in the Pre-pandemic, Pandemic, and Post-pandemic Periods with Moderate-to-Severe Acute Hypoxic Respiratory Failure

|  | Pre-Pandemic<br>1/2018-2/2020<br>(N=1,195) | Pandemic<br>3/2020-2/2022<br>(N=2,969) | Post-Pandemic<br>3/2022-12/2024<br>(N=1,596) |
| --- | --- | --- | --- |
| Age: mean years (SD) | 58.3 (16.2) | 60.3 (14.1) | 58.8 (15.5) |
| Sex: n (% female) | 518 (43.3) | 1,206 (40.6) | 705 (44.2) |
| BMI: mean kg/m<sup>2</sup> (SD) | 31.3 (10.9) | 32.8 (9.9) | 31.5 (11.0) |
| Race/Ethnicity: n (%) |  |  |  |
| Hispanic | 56 (4.9) | 363 (12.2) | 93 (5.8) |
| Non-Hispanic White | 635 (53.1) | 1,366 (46.0) | 836 (52.4) |
| Non-Hispanic Black | 320 (26.8) | 818 (27.6) | 431 (27.0) |
| Non-Hispanic Asian | 45 (3.8) | 164 (5.5) | 79 (4.9) |
| Other | 26 (2.2) | 104 (3.5) | 59 (3.7) |
| Not Reported | 113 (9.5) | 154 (5.2) | 98 (6.1) |
| Non-respiratory SOFA score: mean (SD) | 10 (3) | 9 (3) | 10 (3) |
| FiO<sub>2</sub> at first inclusion timepoint: mean (SD) | 0.9 (.2) | 0.9 (.1) | 0.9 (.2) |
| PEEP at first inclusion timepoint: mean cm H<sub>2</sub>O (SD) | 8 (3) | 11 (4) | 9 (3) |
| Lowest pre-proning PaO<sub>2</sub>/FiO<sub>2</sub> ratio: mean mmHg (SD) | 78 (22) | 80 (22) | 81 (23) |
| PaO<sub>2</sub>/FiO<sub>2</sub> ratio <100: n(%) | 983 (82.3) | 2,258 (76.1) | 1,222 (76.6) |
| First ventilator mode after study enrollment: n (%) |  |  |  |
| Assist/Control Volume Control | 870 (72.8) | 2,258 (76.1) | 1,110 (69.5) |
| Assist/Control Pressure Control | 56 (4.7) | 148 (5.0) | 70 (4.4) |
| Pressure-Regulated Volume Control | 176 (14.7) | 416 (14.0) | 306 (19.2) |
| Other | 76 (6.4) | 88 (3.0) | 68 (4.3) |
| Not reported | 17 (1.4) | 59 (2.0) | 42 (2.6) |
| Time from admission to study enrollment: mean days (SD) | 4.0 (6.2) | 5.2 (6.3) | 5.1 (9.2) |
| Time from intubation to study enrollment: mean days (SD) | 0.8 (0.4) | 0.7 (0.4) | 0.8 (0.4) |
| Vasopressor utilization when met proning criteria: n (%) |  |  |  |
| None | 230 (19.2) | 506 (17.0) | 218 (13.7) |
| ≤0.1 µg/kg/min norepinephrine equivalents | 185 (15.5) | 887 (29.9) | 239 (15.0) |
| >0.1 µg/kg/min norepinephrine equivalents | 780 (65.3) | 1,576 (53.1) | 1,139 (71.4) |
| SARS-CoV2 positivity: n (%) | -- | 1,596 (53.8) | 120 (7.5) |
| Basis for study inclusion: n (%) |  |  |  |
| PROSEVA criteria | 1,169 (97.8) | 2,440 (82.2) | 1,467 (91.9) |
| Prone criteria | 26 (2.2) | 529 (17.8) | 129 (8.1) |
| Treatment at Academic Hospital, n (%) | 982 (82.2) | 1,884 (63.5) | 1,098 (68.8) |
| Hospital ICU capacity (total ICU beds), n (%): |  |  |  |
| Small <20 | 52 (4.4) | 440 (14.8) | 152 (9.5) |
| Medium (20-49) | 164 (13.7) | 623 (21.0) | 335 (21.0) |
| Large (≥50) | 979 (81.9) | 1,906 (64.2) | 1,109 (69.5) |

BMI: body mass index, PEEP: positive end expiratory pressure, SD: standard deviation, SOFA: sequential organ failure assessment

Early initiation of proning occurred in 1,737/5,760 (30.2%) of eligible patients; 96/1,195 (8.0%) pre-pandemic, 1,323/2,969 (44.6%) pandemic, and 318/1,596 (19.9%) post-pandemic (Figure 1). After adjusting for pre-specified covariates, proning use was significantly higher in the pandemic vs. pre-pandemic periods (OR 8.25, 95% CI: 6.35-10.70) and pandemic vs. post-pandemic periods (OR: 2.76, 95% CI: 1.83-4.17; Table 2, Supplement Figure-S2).

**Table 2:** Prone Positioning Use Across Study Periods, SARS-CoV2 Status, and Hospital Type

| Characteristics | Proning in 12 Hours, n/N (%) | Comparison Groups | Adjusted Odds Ratio (95% CI)<sup>a</sup> | P value |
| --- | --- | --- | --- | --- |
| Study Period |  | Primary Analysis |  |  |
| Pre-pandemic | 96/1,195 (8.0) | Pandemic vs Pre-pandemic | 8.25 (6.35-10.70) | <0.001 |
| Pandemic | 1,323/2,969 (44.6) | Reference | - | - |
| Post-pandemic | 318/1,596 (19.9) | Pandemic vs Post-pandemic | 2.76 (1.83-4.17) | <0.001 |
|  |  | Secondary Analyses |  |  |
| SARS-COV2 Status and Period |  |  |  |  |
| Pre-pandemic (no SARS-CoV2) | 96/1,195 (8.0) | Pandemic SARS-CoV2 (-) vs Pre-pandemic | 3.87 (2.92-5.13) | <0.001 |
| Pandemic: SARS-CoV2 + | 965/1,596 (60.5) | Pandemic SARS-CoV2 (+) vs Pandemic SARS-CoV2 (-) | 4.55 (3.85-5.56) | <0.001 |
| Pandemic: SARS-CoV2 - | 358/1,373 (26.1) | Reference | - | - |
| Post-pandemic: SARS-CoV2 + | 48/120 (40.0) | Pandemic SARS-CoV2 (-) vs All post-pandemic | 1.37 (1.03-1.83) | 0.03 |
| Post-pandemic: SARS-CoV2 - | 270/1,476 (18.3) |  |  |  |
|  |  |  | Median Posterior Odds Ratio (95% CrI)<sup>b</sup> |  |
| Hospital Type and Period |  |  |  |  |
| Pre-pandemic: Community | 30/213 (14.1) | Pre-pandemic: community vs academic hospitals | 1.96 (0.72-5.02) |  |
| Pre-pandemic: Academic | 66/982 (6.7) |  |  |  |
| Pandemic: Community | 643/1085 (59.3) | Pandemic: community vs. academic hospitals | 2.28 (1.12-4.14) |  |
| Pandemic: Academic | 680/1884 (36.1) |  |  |  |
| Post-pandemic: Community | 150/498 (30.1) | Post-pandemic: community vs academic hospitals | 2.52 (1.19-5.38) |  |
| Post-pandemic: Academic | 168/1098 (15.3) |  |  |  |

<sup>a</sup>Confidence interval; <sup>b</sup>Credible interval

**Figure 1.**
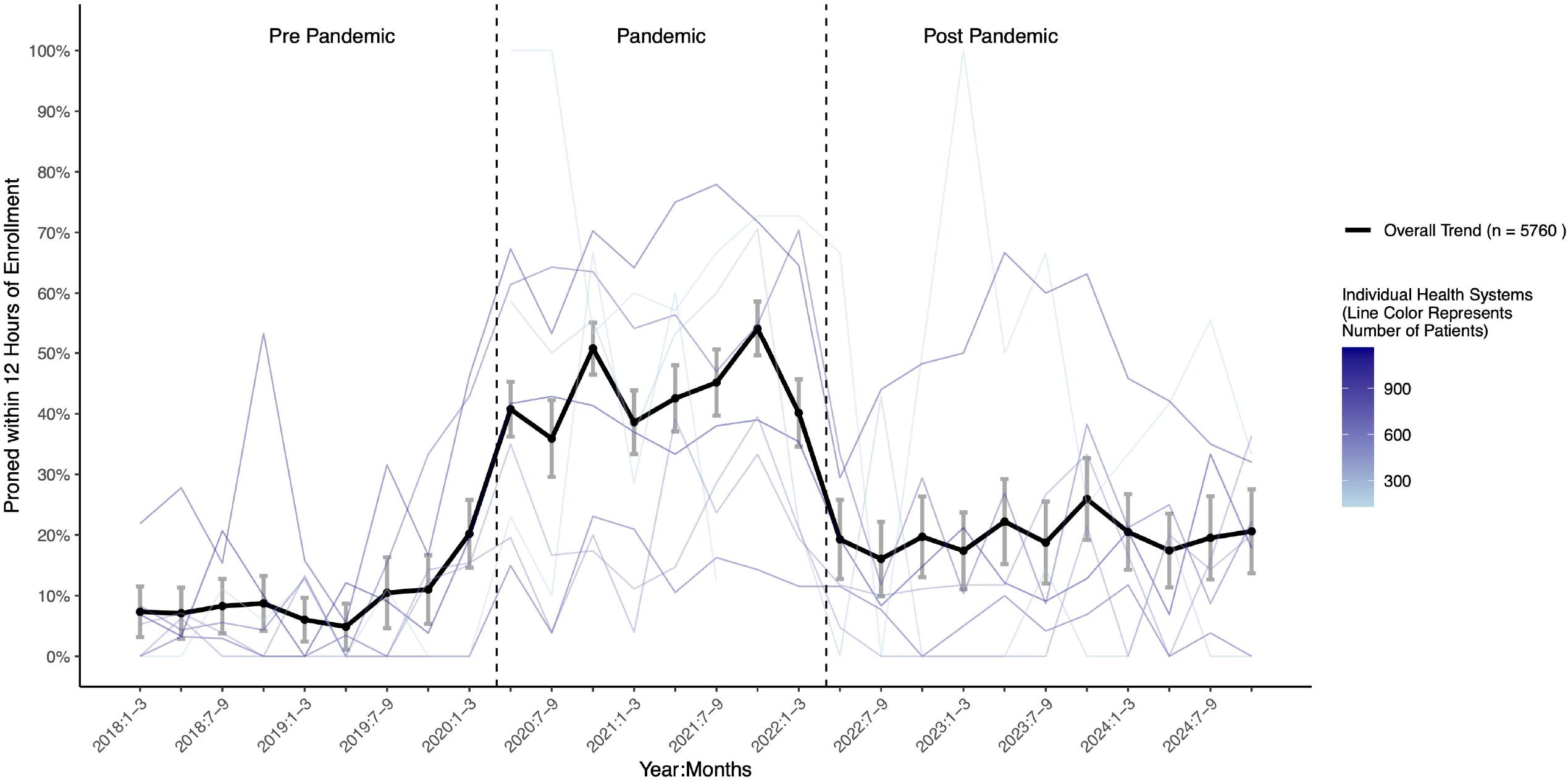
Proning rates across the CLIF Consortium. Variation in proning rates between 1/1/2018-12/31/2024, across nine health systems in the CLIF consortium

In a secondary analysis, pandemic patients with SARS-CoV2 were more likely to be proned than patients without SARS-CoV2 (OR 4.55, [95% CI: 3.85-5.56 Table 2]). Pandemic-period patients without SARS-CoV2 were still more likely to be proned than those admitted pre-pandemic (OR 3.87, [95% CI: 2.92-5.13]) and post-pandemic (OR 1.37, [95% CI: 1.03-1.83]). Consortium-wide trends in proning rates for patients with and without SARS-CoV2 are shown in Figure 2.

**Figure 2.**
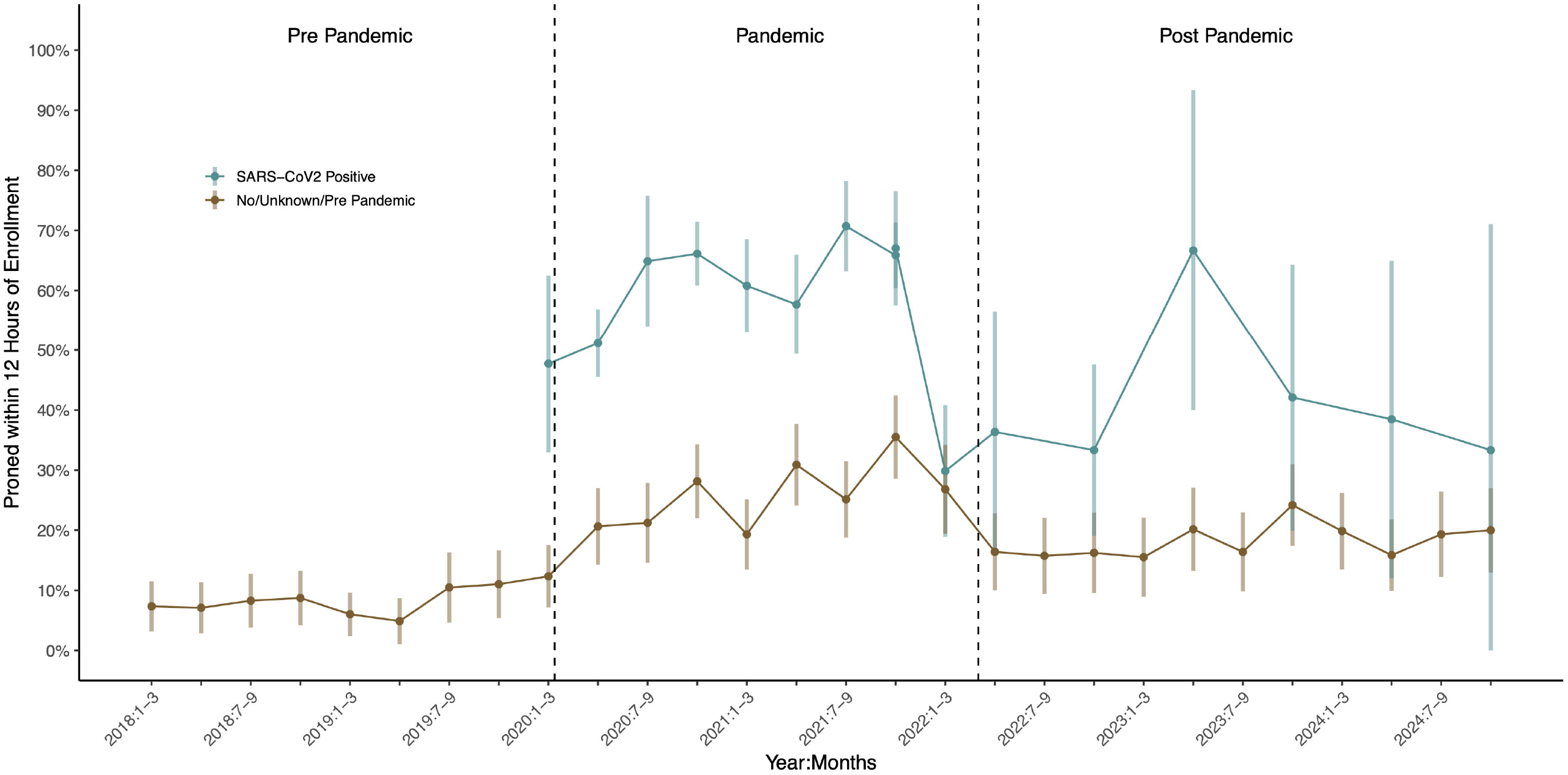
Proning rates by SARS-CoV2 status. Consortium-wide proning rates among patients with and without SARS-CoV2 between 1/1/2018-12/31/2024. Visualized rates reflect average proning rates over three-month periods, with the exception of post-pandemic data for patients positive for SARS-CoV2, which each reflect six months of data given the small number of these patients

Rates of proning by hospital varied considerably in each period, with a median odds ratio (mOR) of 2.54 (95% credible interval [CrI]: 1.75-4.58) pre-pandemic, mOR 2.33 (95% CrI: 1.92-3.04) pandemic, and mOR 2.58 (95% CrI: 1.99-3.73) post-pandemic (Figure 3). Community hospitals were significantly more likely to prone patients than academic hospitals during pandemic and post-pandemic periods, with median posterior ORs of 2.3 (95% CrI: 1.1-4.1) and 2.5 (95% CrI: 1.2-5.4), respectively, but not pre-pandemic (median posterior OR 2.0, 95% CrI: 0.7-5.0; Table 2). There was no difference in proning rates by hospital ICU capacity in any period (Supplement Table-S6).

**Figure 3.**
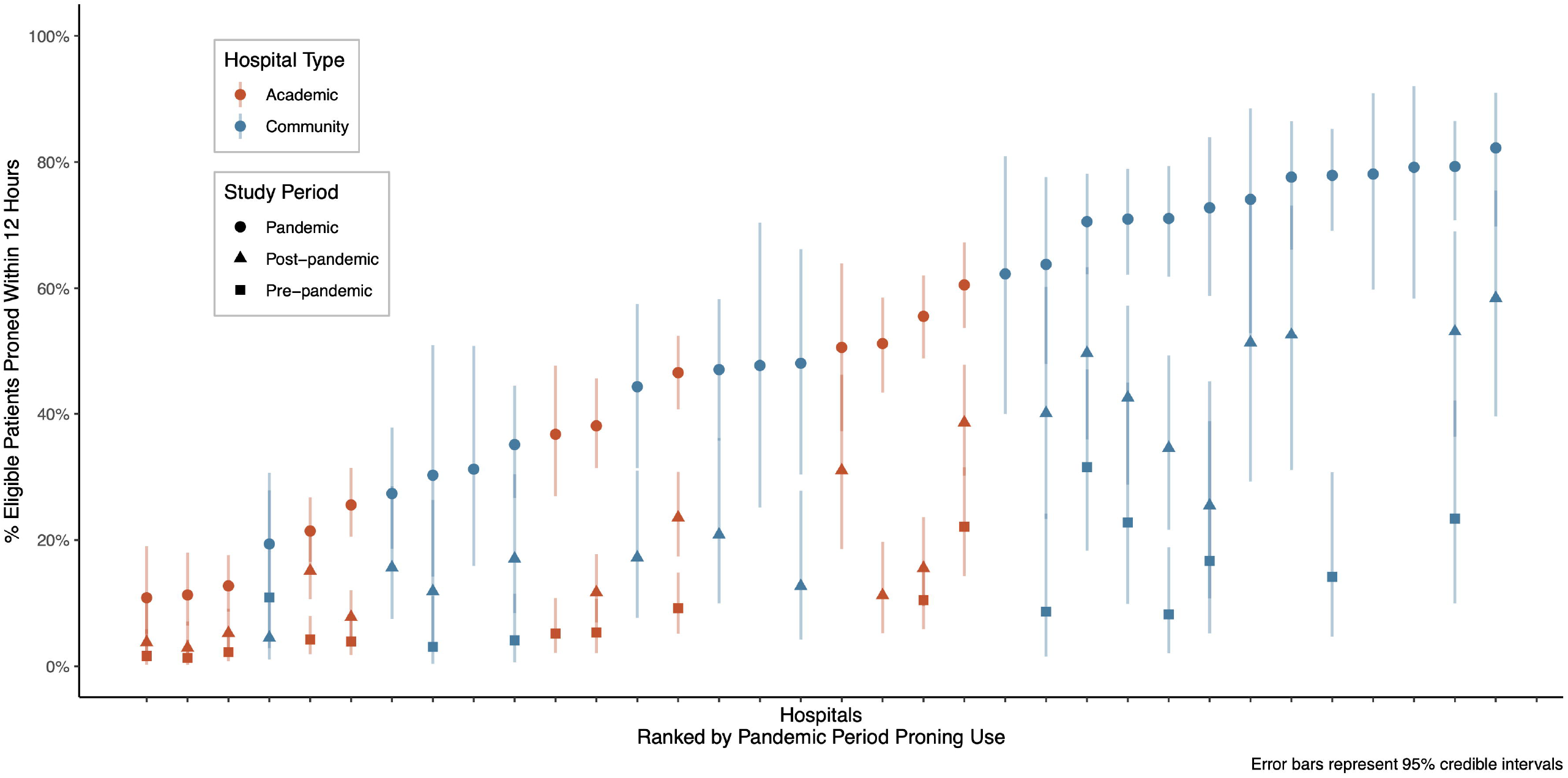
Risk-adjusted hospital-level variation in proning rates across CLIF Consortium. Variation in proning rates between 1/1/2018-12/31/2024, across 34 hospitals with at least 10 patients per period in the CLIF consortium. Hospitals are ranked by pandemic period proning use

The results of the sensitivity analysis evaluating proning rates among the subset of health systems with data across all three periods had nearly identical estimates to the primary analysis (Supplement Table-S7). The results of the sensitivity analyses among patients with severe respiratory failure (PaO_2_/F_i_O_2_ ratio <100) and the analysis that extended timing of early proning events from 12 to 72 hours were both consistent with the primary analysis. In the sensitivity analysis that extended timing to 72 hours, proning rates were 12.8% pre-pandemic, 55.3% pandemic, and 25.2% post-pandemic. Estimates for period associations with proning using Bayesian hierarchical regression were also nearly identical to the primary approach.

## Discussion

In a cohort of patients with moderate-to-severe respiratory failure at 37 North American hospitals, there was an increase in prone positioning during the COVID-19 pandemic followed by a post-pandemic decline. There was considerable hospital-level variability in proning across all periods. During the pandemic, patients with SARS-CoV2 were more likely to be proned than other eligible patients meeting the same hypoxemia criteria, but patients negative for SARS-CoV2 were still more likely to be proned than patients in the pre-pandemic and post-pandemic periods.

This study provides a window into contemporary use of a life-saving ARDS intervention in a large sample with representation from community and academic hospitals. The results confirm small, single-health system findings that suggested proning use declined post-pandemic (5, 7). There are several potential reasons for the decline after increased uptake during the pandemic. First, recognition of ARDS is challenging even among experienced clinicians (14) and evidence-based practices, including prone positioning, are implemented at lower rates when ARDS goes unrecognized (1). ARDS related to SARS-CoV2 may be easier to recognize, as it is more homogeneous. Patients with SARS-CoV2-related ARDS were also often grouped together in the same hospital location during the pandemic, further enhancing ARDS syndromic recognition. In this setting, teams may have developed reflexive responses to prone patients with SARS-CoV2 but not connected these experiences to ARDS from other etiologies. When the incidence of severe COVID-19 dropped in early 2022, SARS-CoV2-related ARDS comprised a smaller proportion of acute hypoxemic respiratory failure, and thus, ARDS recognition may have decreased.

Second, there was an increase in pandemic-era proning even among patients without SARS-CoV2. This may have been due to recency bias, as clinicians accustomed to proning in COVID-19 applied this to ARDS of other etiologies as well (24). The spillover effect found in this study is consistent with other critical care practices, in which pandemic-era practices for patients with SARS-CoV2 affected practice patterns for those without SARS-CoV2 (25). Third, starting early in the pandemic, hospitals invested heavily in proning-related education, protocol development, and proning teams; supports which ended as the pandemic waned (10, 24). Finally, some of the decreases in proning seen here may be due to bias, if the definition of moderate-to-severe persistent hypoxemia more accurately identified patients with SARS-CoV2 as having ARDS than those post-pandemic without SARS-CoV2.

Sustainment of critical care practices is a known challenge across acute care settings (26, 27). The lack of sustained proning use suggests that future implementation strategies must not only promote uptake but also include mechanisms to sustain evidence-based practices over time. One potential strategy for proning is the implementation of a comprehensive ARDS order set, ordered at the time of initial ARDS diagnosis. EHR order sets have been successfully implemented to improve and sustain adherence with other ARDS practices, including low tidal volume ventilation (28). Incorporating proning criteria into an order set of guideline-recommended ARDS practices could reduce the provider burden to sequentially order different treatment strategies as a patient’s ARDS severity worsens.

In this study, the chance of receiving proning was largely determined by which hospital a patient was treated at, even after adjusting for patient and hospital factors. Hospital-level variability is also evident in other ARDS evidence-based practices and such undesired variability in evidence-based practices is a target for implementation interventions (29). Contextual investigations that seek to understand what distinguishes ICUs that frequently deliver proning care and what barriers exist at hospitals that struggle to implement such care are important future steps.

In our cohort, proning was used more commonly at community versus academic hospitals. Rapid initiation of antibiotics in septic shock, another evidence-based, guideline recommended practice, has similarly been shown to occur at higher rates in community hospitals (30). We hypothesize it may be easier to implement and sustain interventions at community hospitals because interventions are directed towards a smaller number of providers that each provide care more frequently. Our finding of improved community hospital proning rates is different than a prior pneumonia-based ARDS study from a single-health system within the CLIF consortium (8). In contrast to that study, our cohort included patients without a pneumonia diagnosis (e.g., trauma and other non-respiratory causes of ARDS), who may be admitted to academic institutions more often and are less likely to be proned (5, 9). Higher proning rates at community hospitals may also reflect differential use of arterial blood gases, a requirement for inclusion in this study. A higher threshold for obtaining an arterial blood gas in community hospitals would mean academic hospitals could include lower acuity patients that are proned less frequently (1, 5, 9).

Strengths of this study include large sample size and inclusion of a diverse set of North American academic and community hospitals, enhancing the generalizability of conclusions. Furthermore, the novel CLIF databases contain granular electronic health record data, which was historically difficult to obtain due to technical limitations and privacy concerns (11). This study applied robust federated methods of statistical analysis to aggregate these data across health systems while maintaining patient privacy. However, there are several limitations. Proning-eligibility was pragmatically defined by persistent moderate-to-severe hypoxemic respiratory failure, rather than clinically adjudicated ARDS. Even with patient adjudication, the diagnosis is difficult for expert clinicians to reliably identify (1, 14) and retrospectively adjudicating patients was not feasible across federated health systems. A single-center study used a similar hypoxemia-based definition of persistent moderate-to-severe respiratory failure and found that over 80% of patients had chest x-ray evidence of ARDS, as adjudicated with a machine learning approach (5). Second, some patients with ARDS may not be included in the study due to local practice variation in arterial blood gas use. Relying on arterial blood gases to meet inclusion criteria may enrich the study cohort with a sicker population, potentially excluding some patients who would benefit from proning. Finally, this study design is unable to distinguish between potential causes of proning changes over time, including higher rates of true ARDS during the pandemic, the effect of recency bias on ARDS diagnosis, and changes in staffing patterns.

## Conclusions

Proning increased during the COVID-19 pandemic, but subsequently declined. The data suggest that use of proning may be largely determined by patient-independent factors such as hospital site, leading to unwanted variation in the use of a mortality-reducing intervention. Interventions that encourage prone positioning in appropriate patients are urgently needed.

## Data Availability

This study is a secondary analysis of electronic health record data
collected at 37 hospitals as part of routine patient care. While these individual patient data will not be made available, the statistical code for cohort generation and analysis are available on github (https://github.com/Common-Longitudinal-ICU-dataFormat/CLIF_Proning_Incidence_Severe_ARF.git).

https://github.com/Common-Longitudinal-ICU-data-Format/CLIF_Proning_Incidence_Severe_ARF.git

## Acknowledgements

Sources of Funding: AKB: NIH/NHLBI T32HL007749; PGL: NIH/NCI K08CA270383, NIH UL1TR002369; CAG: NIH/NHLBI K23HL169815, Francis Family Foundation Parker B. Francis Opportunity Award, American Thoracic Society Unrestricted Grant; ACO: NIH/NHLBI T32HL007891, JCR: NIH/NIDA R01DA051464; WFP: NIH R01LM014263; MWS: NIH/NHLBI R01HL158626, CHH: NIH/NHLBI K23HL169743.

## Supplemental Material Content

Table S1. Health System Characteristics and IRB Exemptions/Approvals.

Table S2. Strengthening the Reporting of Observational Studies in Epidemiology Statement Checklist

Table S3. Prone Positioning Documentation Table S4. Missing Covariate Data

Table S5. Priors for Bayesian Analyses

Figure S1. Distribution of SARS-CoV2 Positive and Negative Patients

Figure S2. Meta Analysis of Primary Outcome

Table S6: Prone Postioning Use Across Study Periods and Hospital Size

Table S7: Sensitivity Analyses Showing Period Effect Estimates

