## Supplemental Material for "Prone Positioning in a North American Cohort of Hypoxemic Patients on Mechanical Ventilation"

**Table S1: Health System Characteristics and IRB Exemptions/Approvals.**

| Health System | Location | Period of study data | Hospitals | IRB study number | IRB Status |
| --- | --- | --- | --- | --- | --- |
| University of Pennsylvania | East coast | 1/1/2018-12/31/2024 | 3 academic  3 community | 853717 | Approved |
| Johns Hopkins University | East coast | 1/1/2018-12/31/2024 | 2 academic  3 community | IRB0042173 | Exempt |
| University of Michigan | Midwest | 1/1/2018-12/31/2024 | 1 academic | HUM00260807 | Exempt |
| Northwestern University | Midwest | 1/1/2018-12/31/2024 | 1 academic  8 community | STU00202840 | Approved |
| University of Minnesota | Midwest | 1/1/2018-12/31/2024 | 1 academic  11 community | STUDY00014815 | Approved |
| Rush University | Midwest | 3/1/2020-12/31/2024 | 1 academic | 24082804-IRB | Approved |
| University of Chicago | Midwest | 1/1/2018-12/31/2024 | 1 academic | IRB20-1823 | Approved |
| Oregon Health & Science University | West coast | 3/1/2020-12/31/2024 | 1 academic  1 community | 00025188 | Approved |
| Sunnybrook Health Sciences Centre | Canadian | 1/1/2018-9/30/2021 | 1 academic | 3690 | Approved |

IRB: Institutional review board

**Table S2: Strengthening the Reporting of Observational Studies in Epidemiology Statement Checklist**^1^

|  | Item No | Recommendation | Paper |
| --- | --- | --- | --- |
| **Title and abstract** | 1 | (*a*) Indicate the study’s design with a commonly used term in the title or the abstract | (a) See abstract |
|  |  | (*b*) Provide in the abstract an informative and balanced summary of what was done and what was found | (b) See abstract |
| Introduction | | |  |
| Background/rationale | 2 | Explain the scientific background and rationale for the investigation being reported | See introduction: paragraphs 1-2 |
| Objectives | 3 | State specific objectives, including any prespecified hypotheses | See introduction: paragraph 3 |
| Methods | | |  |
| Study design | 4 | Present key elements of study design early in the paper | See methods: data source |
| Setting | 5 | Describe the setting, locations, and relevant dates, including periods of recruitment, exposure, follow-up, and data collection | See methods: data source, study population |
| Participants | 6 | (*a*) Give the eligibility criteria, and the sources and methods of selection of participants. Describe methods of follow-up | See methods: study population |
|  |  | (*b*) For matched studies, give matching criteria and number of exposed and unexposed | N/A |
| Variables | 7 | Clearly define all outcomes, exposures, predictors, potential confounders, and effect modifiers. Give diagnostic criteria, if applicable | See methods: exposures and outcomes, patient and hospital-level covariates |
| Data sources/ measurement | 8* | For each variable of interest, give sources of data and details of methods of assessment (measurement). Describe comparability of assessment methods if there is more than one group | See methods: exposures and outcomes |
| Bias | 9 | Describe any efforts to address potential sources of bias | See methods: statistical analysis |
| Study size | 10 | Explain how the study size was arrived at | See methods: study population |
| Quantitative variables | 11 | Explain how quantitative variables were handled in the analyses. If applicable, describe which groupings were chosen and why | See methods: statistical analysis |
| Statistical methods | 12 | (*a*) Describe all statistical methods, including those used to control for confounding | See methods: statistical analysis |
|  |  | (*b*) Describe any methods used to examine subgroups and interactions | See methods: statistical analysis |
|  |  | (*c*) Explain how missing data were addressed | See methods: statistical analysis |
|  |  | (*d*) If applicable, explain how loss to follow-up was addressed | N/A |
|  |  | (*e*) Describe any sensitivity analyses | See methods: statistical analysis |
| **Results** | | | |
| Participants | 13* | (a) Report numbers of individuals at each stage of study—eg numbers potentially eligible, examined for eligibility, confirmed eligible, included in the study, completing follow-up, and analyzed | See results: paragraph 1 |
|  |  | (b) Give reasons for non-participation at each stage | N/A |
|  |  | (c) Consider use of a flow diagram | N/A |
| Descriptive data | 14* | (a) Give characteristics of study participants (eg demographic, clinical, social) and information on exposures and potential confounders | See results: paragraph 1 |
|  |  | (b) Indicate number of participants with missing data for each variable of interest | eTable 4 |
|  |  | (c) Summarize follow-up time (eg, average and total amount) | N/A |
| Outcome data | 15* | Report numbers of outcome events or summary measures over time | See results: paragraph 2 |
| Main results | 16 | (*a*) Give unadjusted estimates and, if applicable, confounder-adjusted estimates and their precision (eg, 95% confidence interval). Make clear which confounders were adjusted for and why they were included | See results: paragraph 2; see methods: statistical analysis |
|  |  | (*b*) Report category boundaries when continuous variables were categorized | See methods: statistical analysis |
|  |  | (*c*) If relevant, consider translating estimates of relative risk into absolute risk for a meaningful time period | See results: paragraph 2 |
| Other analyses | 17 | Report other analyses done—eg analyses of subgroups and interactions, and sensitivity analyses | See results: paragraph 3-5 |
|  | **Item No** | **Recommendation** | **Paper** |
| Discussion | | |  |
| Key results | 18 | Summarize key results with reference to study objectives | See discussion: paragraph 1 |
| Limitations | 19 | Discuss limitations of the study, taking into account sources of potential bias or imprecision. Discuss both direction and magnitude of any potential bias | See discussion: paragraph 3, 7-8 |
| Interpretation | 20 | Give a cautious overall interpretation of results considering objectives, limitations, multiplicity of analyses, results from similar studies, and other relevant evidence | See discussion: paragraph 1-8 |
| Generalizability | 21 | Discuss the generalizability (external validity) of the study results | See discussion: paragraph 7 |
| Other information | | |  |
| Funding | 22 | Give the source of funding and the role of the funders for the present study and, if applicable, for the original study on which the present article is based | See article information |

*Give information separately for exposed and unexposed groups.

**Table S3: Prone Positioning Documentation**

| Documented Position | Number of Events |
| --- | --- |
| prone | 53692 |
| prone;pillow support | 7596 |
| prone;head repositioned, right | 5658 |
| prone;head repositioned, left | 5437 |
| prone;lying left side | 2476 |
| prone;reverse trendelenburg | 1997 |
| prone;lying right side | 1728 |
| turned;prone | 1669 |
| turns self;prone | 1578 |
| prone-swimmers left | 1573 |
| prone-swimmers right | 1557 |
| prone;turned right side- at least 30 degrees | 1273 |
| prone;turned left side- at least 30 degrees | 1193 |
| prone;other (comment) | 1184 |
| turned;prone;head repositioned, left | 1068 |
| turned;prone;head repositioned, right | 1053 |
| prone position | 883 |
| prone;turned;right | 682 |
| prone;turned;left | 641 |
| prone;head right | 639 |
| prone;head left | 614 |
| Prone-foam dressings to pressure points | 608 |
| 15 degree micro-turn (icu only);prone | 587 |
| head left;prone;reverse trendelenburg | 537 |
| head right;prone;reverse trendelenburg | 519 |

Twenty-five most common case insensitive strings identified in flowsheet position documentation across the CLIF consortium. Search strings used to identify prone positioning varied by site, including case insensitive “prone” and “swim”.
**Table S4: Missing Covariate Data**

| Variable | HS1 | HS2 | HS3 | HS4 | HS5 | HS6 | HS7 | HS8 | HS9 |
| --- | --- | --- | --- | --- | --- | --- | --- | --- | --- |
| Time period | 0.0% | 0.0% | 0.0% | 0.0% | 0.0% | 0.0% | 0.0% | 0.0% | 0.0% |
| Age | 0.0% | 0.0% | 0.0% | 0.0% | 0.0% | 0.0% | 0.0% | 0.0% | 0.0% |
| Sex | 0.0% | 0.0% | 0.0% | 0.0% | 0.0% | 0.0% | 0.0% | 0.0% | 0.0% |
| Body mass index | 0.0% | 0.6% | 0.0% | 2.0% | 1.7% | 0.5% | 8.4% | 75.7%; 13.3% after multiple imputation of weight | 2.1% |
| Non-respiratory SOFA score | 0.0% | 0.2% | 0.0% | 0.0% | 0.0% | 6.1% | 0.0% | 0.7% | 0.7% |
| PaO_2_/F_i_O_2_ ratio | 0.0% | 0.0% | 0.0% | 0.0% | 0.0% | 0.0% | 0.0% | 0.0% | 0.0% |
| Vasopressor utilization | 0.0% | 0.0% | 0.0% | 0.0% | 0.0% | 0.0% | 0.0% | 0.0% | 0.0% |

HS8 was not permitted to collect race and ethnicity data per health system policy. Thus, 100% of race and ethnicity data in this health system was unreported. Race and ethnicity data were not included as covariates in the pre-specified regression models. HS: health system

**Table S5: Priors for Bayesian Analyses**

| **Variable in Bayesian Model** | **Prior Description** | **Prior Implementation**  **Distribution Type (Mean, SD)** |
| --- | --- | --- |
| Pre-Pandemic Period Proning | Weakly informative. Centered around probability of proning of 12% with 95% CI of 5-50% | Normal (-2, 1) |
| Pandemic Period Proning | Weakly informative. Centered around probability of proning of 50% with 95% CI of 12-88% | Normal (0, 1) |
| Post-Pandemic Period Proning | Weakly informative. Centered around probability of proning of 12% with 95% CI of 5-50% | Normal (-2, 1) |
| Hospital Type (Community vs Academic) | Flat prior. Log odds centered at 0 with a 95% CI of -2 to 2. | Normal (0,1) |
| Hospital Intensive Care Unit Size (Small vs Medium vs Large) | Flat prior. Log odds centered at 0 with a 95% CI of -2 to 2. | Normal (0,1) |

**Figure S1: Distribution of SARS-CoV2 Positive and Negative Patients**

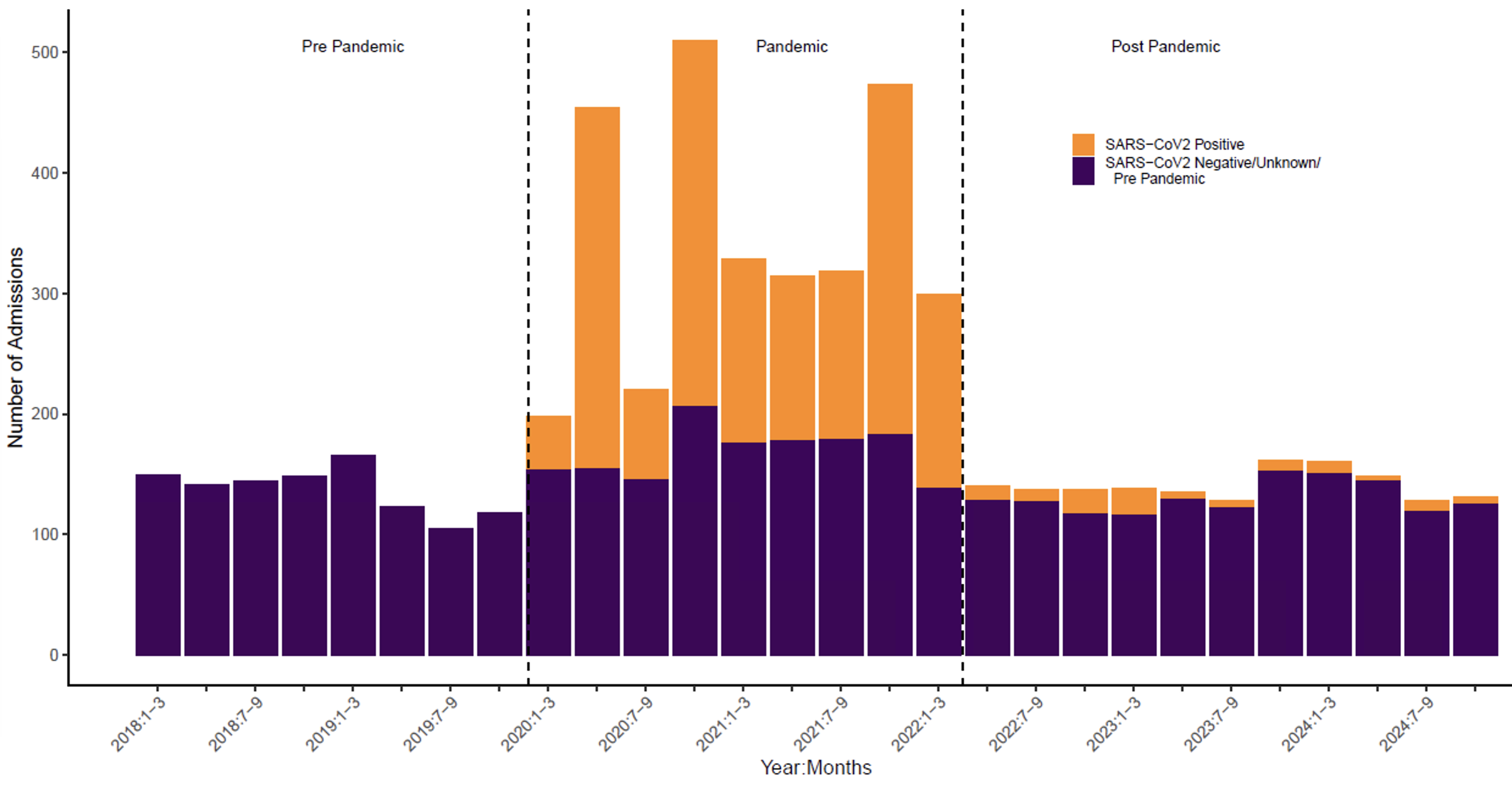

Cumulative SARS-CoV2 positive and negative admissions across the CLIF consortium during serial three-month time periods from 1/1/2018-12/31/2024

**Figure S2: Meta Analysis of Primary Outcome**

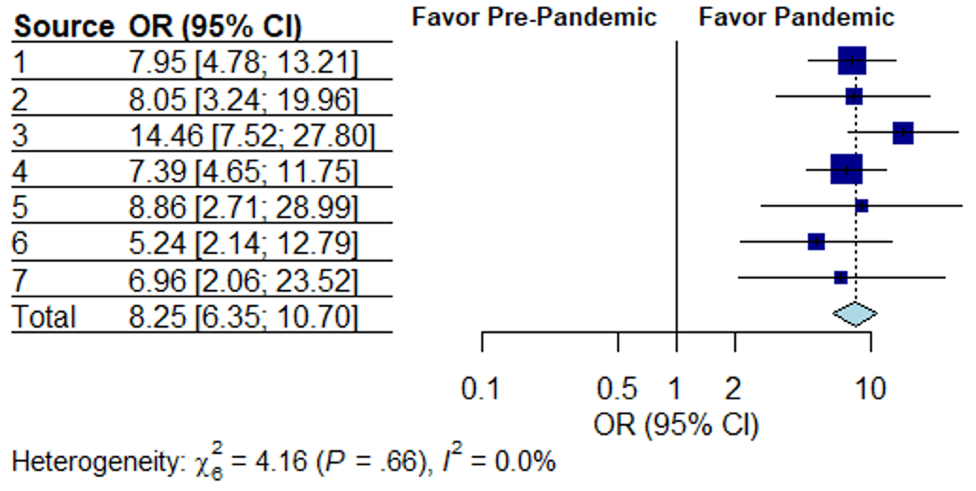
a)

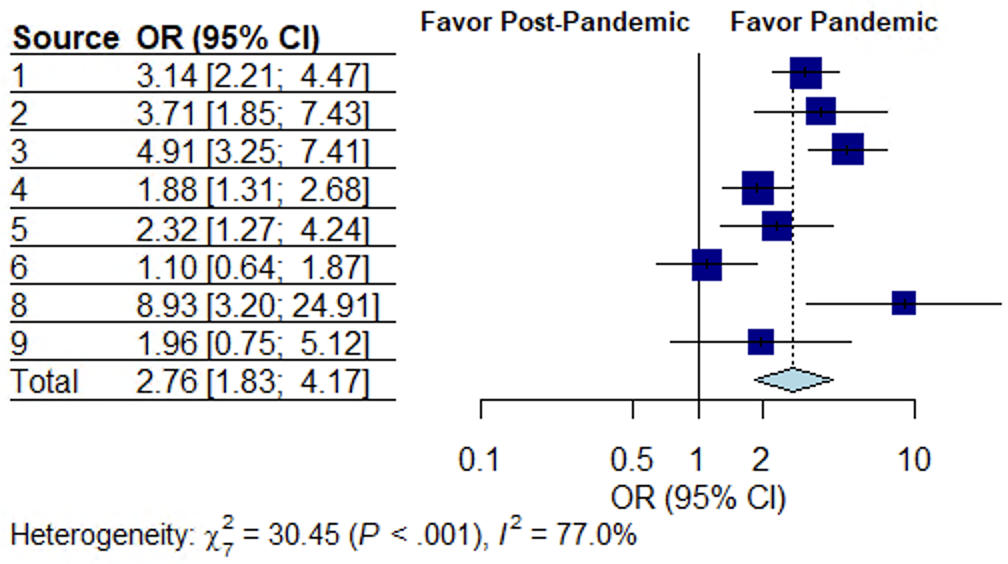

b)

Aggregate individual health system estimates for the odds ratio of proning comparing the pandemic period with the A) pre-pandemic and B) post-pandemic period.

**Table S6: Prone Positioning Use Across Study Periods and Hospital Size**

| **Hospital Intensive Care Unit Size and Period** | **Median Posterior Odds Ratio (95% CrI)**^a^ |
| --- | --- |
| **Pre-pandemic** |  |
| Medium vs Small | 2.10 (0.51-8.59) |
| Large vs Small | 1.15 (0.31-4.45) |
| **Pandemic** |  |
| Medium vs Small | 0.61 (0.29-1.28) |
| Large vs Small | 0.56 (0.29-1.19) |
| **Post-pandemic** |  |
| Medium vs Small | 0.68 (0.25-1.94) |
| Large vs Small | 0.53 (0.21-1.45) |

^a^Credible interval

**Table S7: Sensitivity Analyses Showing Period Effect Estimates**

| Sensitivity Analysis | Adjusted Odds Ratio (95% CI) | P-Value |
| --- | --- | --- |
| **Sensitivity Analyses** | | |
| **Health systems with data across all 3 periods** |  |  |
| Pandemic vs Pre-pandemic | 8.31 (6.37-10.86) | <0.001 |
| Pandemic vs Post-pandemic | 2.55 (1.66-3.91) | <0.001 |
| **Severe hypoxemic respiratory failure (PaO_2_/F_i_O_2_ <100)** |  |  |
| Pandemic vs Pre-pandemic | 7.97 (5.98-10.62) | <0.001 |
| Pandemic vs Post-pandemic | 2.46 (1.63-3.71) | <0.001 |
| **72-hour proning eligibility** |  |  |
| Pandemic vs Pre-pandemic | 7.34 (5.32-10.11) | <0.001 |
| Pandemic vs Post-pandemic | 3.22 (2.17-4.79) | <0.001 |
| **Bayesian Hierarchical Regression** | | |
| Pandemic vs Pre-pandemic | 8.17 (5.30-14.10) | -- |
| Pandemic vs Post-pandemic | 2.80 (2.09-3.86) | -- |
